# Radius of Gyration as predictor of COVID-19 deaths trend with three-weeks offset

**DOI:** 10.1101/2021.01.30.21250708

**Authors:** Alberto Hernando, David Mateo, Jordi Bayer, Ignacio Barrios

## Abstract

Total and perimetral lockdowns were the strongest nonpharmaceutical interventions to fight against Covid-19, as well as the with the strongest socioeconomic collateral effects. Lacking a metric to predict the effect of lockdowns in the spreading of COVID-19, authorities and decision-makers opted for preventive measures that showed either too strong or not strong enough after a period of two to three weeks, once data about hospitalizations and deaths was available. We present here the *radius of gyration* as a candidate predictor of the trend in deaths by COVID-19 with an offset of three weeks. Indeed, the radius of gyration aggregates the most relevant microscopic aspects of human mobility into a macroscopic value, very sensitive to temporary trends and local effects, such as lockdowns and mobility restrictions. We use mobile phone data of more than 13 million users in Spain during a period of one year (from January 6^th^ 2020 to January 10^th^ 2021) to compute the users’ daily radius of gyration and compare the median value of the population with the evolution of COVID-19 deaths: we find that for all weeks where the radius of gyration is above a critical value (70% of its pre-pandemic score) the number of weekly deaths increases three weeks after. The reverse also stands: for all weeks where the radius of gyration is below the critical value, the number of weekly deaths decreased after three weeks. This observation leads to two conclusions: i) the radius of gyration can be used as a predictor of COVID-19-related deaths; and ii) partial mobility restrictions are as effective as a total lockdown as far the radius of gyration is below this critical value.

**Background:** Authorities around the World have used lockdowns and partial mobility restrictions as major nonpharmaceutical interventions to control the expansion of COVID-19. While effective, the efficiency of these measures on the number of COVID-19 cases and deaths is difficult to quantify, severely limiting the feedback that can be used to tune the intensity of these measures. In addition, collateral socioeconomic effects challenge the overall effectiveness of lockdowns in the long term, and the degree by which they are followed can be difficult to estimate. It is desirable to find both a metric to accurately monitor the mobility restrictions and a predictor of their effectiveness.

**Methods:** We correlate the median of the daily radius of gyration of more than 13M users in Spain during all of 2020 with the evolution of COVID-19 deaths for the same period. Mobility data is obtained from mobile phone metadata from one of the major operators in the country.

**Results:** The radius of gyration is a predictor of the trend in the number of COVID-19 deaths with 3 weeks offset. When the radius is above/below a critical threshold (70% of the pre-pandemic score), the number of deaths increases/decreases three weeks later.

**Conclusions:** The radius of gyration can be used to monitor in real time the effectiveness of the mobility restrictions. The existence of a critical threshold suggest that partial lockdowns can be as efficient as total lockdowns, while reducing their socioeconomic impact. The mechanism behind the critical value is still unknow, and more research is needed.

## 1. Introduction

The World Health Organization (WHO) declared the ongoing COVID-19 international crisis as a pandemic on March 11, 2020 [1]. To that date, over 118,000 positive cases had been reported in over 110 countries and territories around the world. The spread risk became real as the number of cumulated confirmed cases grew to 31.7 million in October Worlwide, with more than 1.1 million deceases [2].

The first positive case of COVID-19 in the Spanish territory was detected on January 31, 2020 [3]. It was followed by many others in the following weeks. As of March 9, the number of cases exploded in Madrid, forcing authorities to impose partial mobility restrictions. However, positive cases had already been detected in all the regions of Spain and the government declared a national lockdown with so called State of Alarm on March 15th [4]. Many other countries adopted similar measures, with strong restrictions in mobility that proved to be efficient to control the spreading of the virus [5-8]. However, they ultimately led to multiple economic and social collateral effects [9].

A recent study showed that the centralized structure of the country’s communication network contributed to exporting positive cases from Madrid to all other Spanish provinces [10]. Indeed, the number of visits per capita from travellers between Madrid and a given province in the weeks before the lockdown strongly correlates with the peak of sickness incidence, mortality, and antibody prevalence in the first pandemic wave. Three months of lockdown with a strict reduction in both intra- and inter-city mobility helped reduce the number of new cases to a minimum. However, a rebound in the number of cases in summer put the country in alert again---weeks earlier than the rest of Western Europe---potentially due to a premature or abrupt easing of the lockdown [11]. A second State of Alarm was declared at the end of October by the Spanish government and new regional mobility restrictions lead to a stabilization of the crisis, but its lifting in December and ultimately during Christmas period led to a high peak in the number of cases a few weeks later [1].

The central role of human mobility in epidemics has been extensively considered in the past [13-20] and more recently for COVID-19 in particular [11,21,22]. It is well known that mobility patterns correlate with the socio-economic structure of cities. Urban space segregation modulates the social opportunities of its inhabitants and may generate inequalities of access to resources and jobs [23-25]. While these inequalities have been reported in the past, it is still unknown how the pandemic will affect the social equilibrium in the post-pandemic era, the so-called *new normal*.

The explicit correlation between mobility and the spread of COVID-19 in Spain was obtained after analysing Call Detail Records (CDR) data from a major mobile operator. CDR data has been widely used to understand mobility in general [26,27] and recent studies are showing its convenience for tracking the correlations with the pandemic effects in particular [28,29]. The radius of gyration is a useful metric to quantify the mobility of a region by place of residence which has been show also useful to measure the effects of the lockdown in the pandemic [22,28,30]. This metric was recently used to measure the effects of the lockdown and the evolution of the epidemic in groups of population segmented by the average salary, finding remarkable differences between low- and high-income populations and its potential role in the second wave during the Spanish summer and the expansion of COVID-19 in Europe [31].

In view of the potential of the above cited tools to understand the effects of the pandemic, we present in this work measurements of the radius of gyration per person for all Spain in 2020 to find potential correlations with the number of hospitalizations and deaths by Covid-19 along 2020 and beginning of 2021. To this aim, we use CDR data from 13 million users homogeneously distributed in the Spanish territory, which represent one of the largest datasets ever reported. We find that the changes in radius of gyration accurately reflect the lockdowns and partial mobility restrictions taken place during 2020. In addition, we find a remarkable correlation with COVID-19 hospitalizations and deaths: for all weeks where the radius of gyration is above a critical threshold (70% of its pre-pandemic score) the number of new weekly hospitalizations increased after two weeks, and the number of weekly deaths after three; for all weeks where the radius of gyration is below the critical threshold, the number of new weekly hospitalizations decreased after two weeks, and the number of weekly deaths decreased after three.

In brief, our results demonstrate that i) planning of nonpharmaceutical interventions to COVID-19 such as lockdowns and other mobility restrictions can benefit from tracking the radius of gyration as a predictor of effectiveness in fighting the spread of the pandemic; and ii) partial restrictions can control the pandemic without the need of total lockdowns as long they lower the radius of gyration below its critical, thus reducing the collateral socioeconomic effects of total lockdowns.

## 2. Data and Methods

We use data on COVID-19 hospitalizations and deaths downloaded from the COVID-19 Panel [32], obtained from the recorded cases by the National Epidemiologic Watcher (RENAVE) through the Web SiViES platform (Spanish Watch System) managed by the National Epidemiologic Centre (CNE).

Mobility is measured using Call Detail Records from one of the major telecommunication operators in Spain. The data set includes 13 million users in a timespan of 42 weeks, from 10th of January 2020 to 18th of October 2024. A user’s residence is defined as the most common census section visited from 10pm to 6am during a given month. The sample is scaled per each census section to reproduce the total population as reported by INE. The daily radius of gyration for a user is measured as [27]

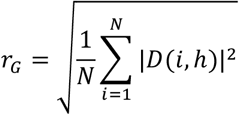

where *N* is the number of events per each user and *D*(*i, h*) is the geodesic distance between the i-th daily event and the user’s residence. We assign a weekly value per user as the weekly average (7 days from Monday to Sunday). We obtain the long-tailed distribution depicted in Fig. 1 when aggregating for all users (pre-pandemic). The mode of the distribution is located at the lowest bin of 1 km with a median of 3.2 km and a mean value of 8.0 km. The big difference between median and mean is indeed typical of these long-tailed distributions. In the following, we will use the median per week as representative statistic of the distribution of daily radiuses of gyration of the entire country.

**Fig. 1.**
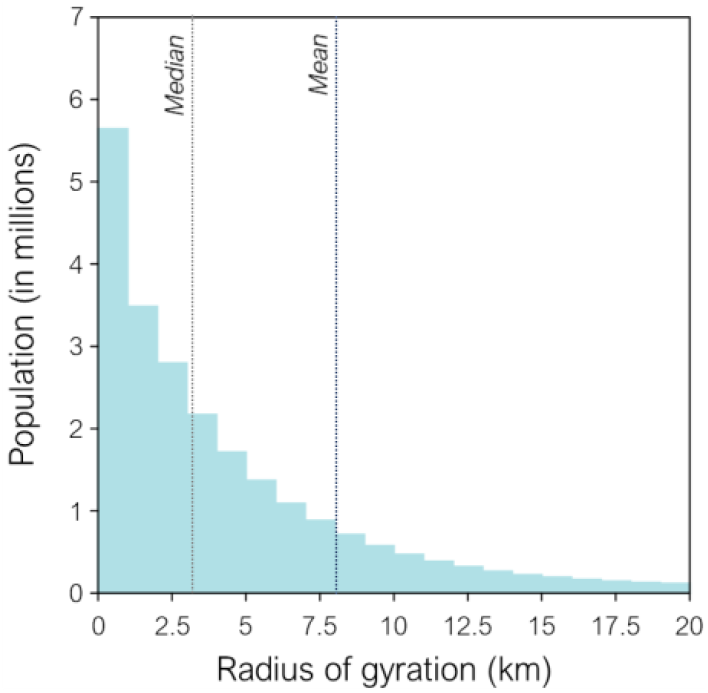
Distribution of radius of gyration r_G_ (pre-pandemic) for all Spanish population. *Important note*: estimating the radius of gyration from mobile phone data is a non-trivial task, as i) the frequency of network events does not reflect the relevance of those events and ii) it is very sensitive to the algorithm used for users’ residence. A calibrated and tested method for computing *r*_*G*_ is presented at the Supplementary Information.

## 3. Results

### 3.1. Median of radius of gyration as a measure of national mobility

The radius of gyration, computed as described in Supplementary Information, reflects the effects of the restrictions in mobility as well as the underlying trends in people’s behaviour week per week, as shown in Fig. 2. Being *R*_*G*_(*t*) the median radius of gyration for week *t*, we can recognize up to 10 periods correlated with the implemented measures and special national dates along 2020 (for a detailed summary of the COVID-19 chronology in Spain see Ref. [33]):

**Fig. 2.**
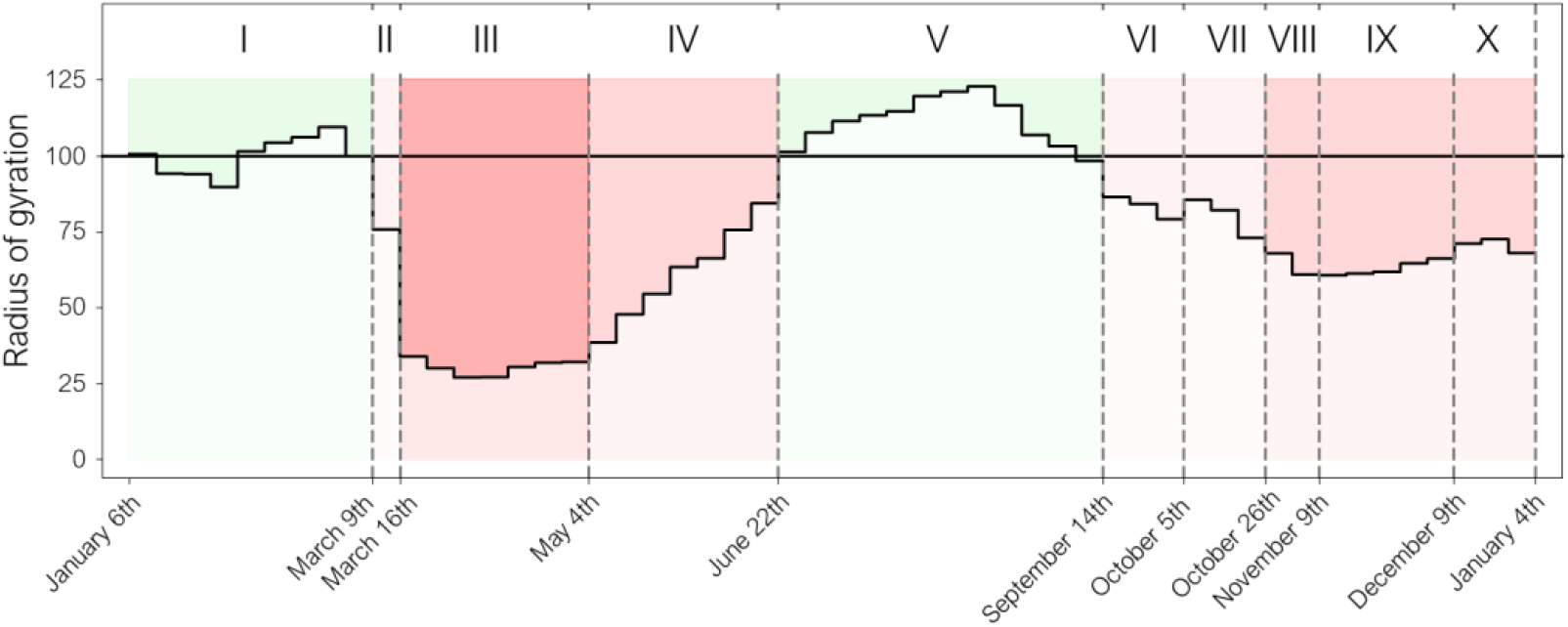
Evolution of median radius of gyration *R*_*G*_(*t*) along 2020 (see text). *I. Pre-pandemic*. The mean value of *R*_*G*_ in the first weeks before the state of alarm is 3.2 km. No restrictions were yet adopted in this period and we will use this value as reference for the rest of the year. *II. First restrictions*. The Community of Madrid was the first Spanish region to adopt mobility restrictions, one week before the rest of the country. *III. First state of alarm*. The Spanish government declared the state of alarm, imposing a total national lockdown which reduced *R*_*G*_ down to a 25% of its pre-pandemic value. *IV. Easing lockdown*. The Spanish government defined several phases in the easing adapted to the regional situation of the epidemic. *R*_*G*_ grows as the different regions transited to the so-called *New Normal*, reaching pre-pandemic values. *V. Summer holidays*. With a very low number of new COVID-19 hospitalizations and deaths, no major mobility restrictions are in effect at the national level, allowing citizens to plan their summer holidays. National tourism expanded rapidly, with a peak in August, as reported by specialized local journals [34]. This period sees an increase of *R*_*G*_ up to a 125% to its pre-pandemic value. *VI. Partial restrictions in the New Normal*. The new normal is characterized by, among other factors, telecommuting (TC) and perimetral lockdowns. We can see these effects as a general decrease of *R*_*G*_ down to 80%. *VII. National Day of Spain of October 12*^*th*^. Regional authorities adopted special measures to limit the mobility during the large weekend of the National Day of Spain. However, these measures failed in preventing massive displacements [35,36] reflected as an increase in *R*_*G*_ breaking its downward trend of the previous weeks. *VIII. Second state of alarm*. The Spanish government declared a second state of alarm to prevent massive displacements during the national holiday of November 1^st^, which successfully reduced *R*_*G*_ to its minimum value since the total lockdown, at 60%. *IX. Partial restrictions*. Strategic perimetral lockdowns and curfews followed the second state of alarm, leading to a smooth increase of national *R*_*G*_. *X. Christmas holidays*. Despite the pandemic and the local mobility restrictions, *R*_*G*_ peaked beyond 70% in the previous days and during Christmas holidays, decreasing only after the New Year’s Eve.

### 3.2. Radius of gyration VS COVID-19 deaths

We define the critical value 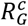 and the offset time *t*_*c*_ as the solutions of the equation

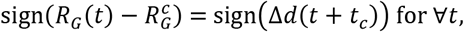

where *d*(*t*) is the number of COVID-19 deaths for week *t* and Δ*d*(*t*) is the increment of *d*(*t*) between consecutive weeks, numerically obtained as Δ*d*(*t*) = *d*(*t*) − *d*(*t* − 1). This equation is fulfilled if when *R*_*G*_(*t*) is *above* 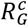, then Δ*d*(*t*) is *positive* and thus *d*(*t*) *increases* after *t*_*c*_ weeks, while when *R*_*G*_ is *below* 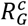, then Δ*d*(*t*) is *negative* and thus *d*(*t*) *increases* after *t*_*c*_ weeks.

This equation may not have a solution for all *t*, but it may show an optimal solution for the largest range of values of *t*. A practical metric to evaluate the fitness of the solution is the Pearson’s correlation as

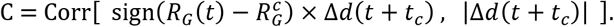

Indeed, if 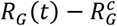and Δ*d*(*t* + *t*_*c*_) are in phase, C = 1; if both functions and in opposite phase, C = −1; and if their phase is randomly distributed, C = 0. This correlation also presents a desired property as it gives more weight to those weeks with high value of Δ*d*, while gives less weight to weeks with small values of Δ*d*. After exploring for several values of 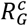 and *t*_*c*_, we found as optimal solution (see Fig. 3):

**Fig. 3.**
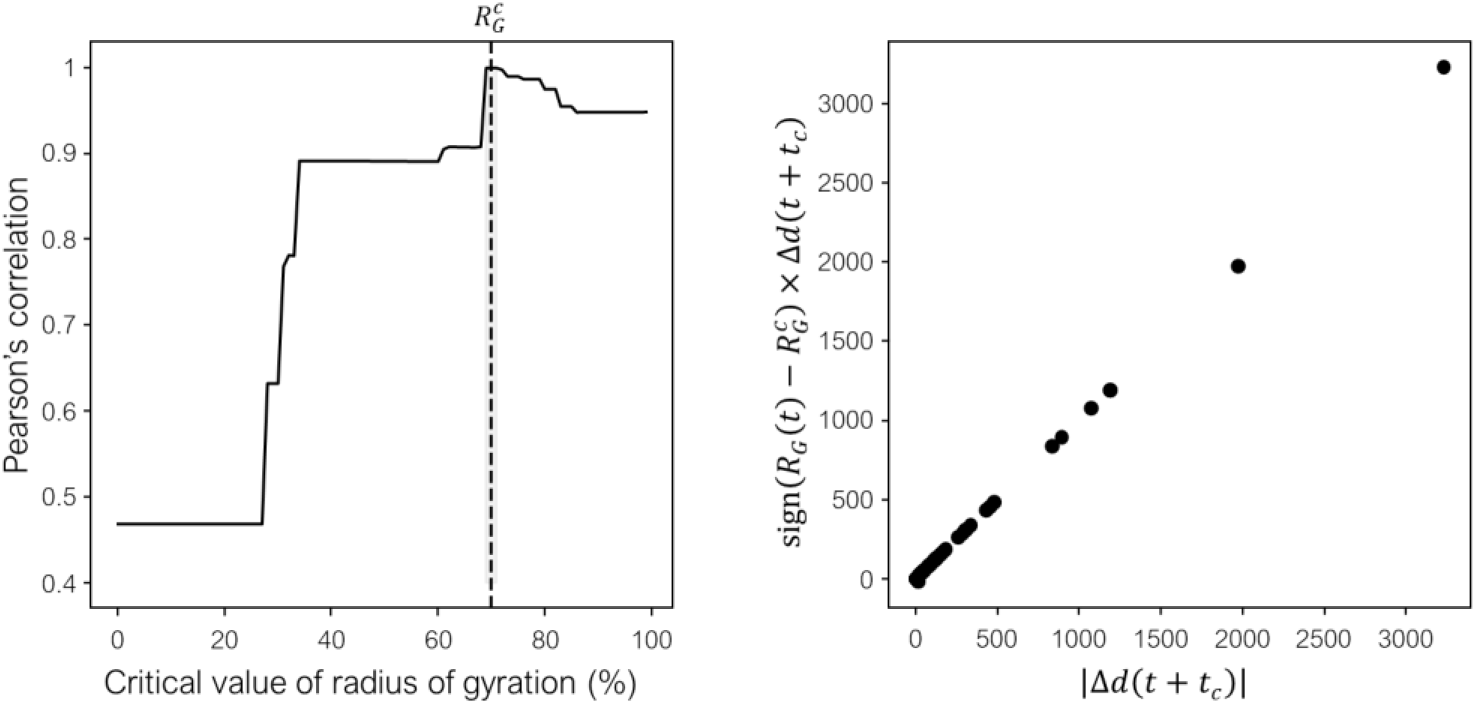
Left: Pearson’s correlation for the optimal value *t*_*c*_ = 3 weeks for different values of 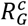 with respect the pre-pandemic value. Right: 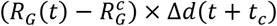 versus |Δ*d*(*t* + *t*_*c*_)| at optimal values.

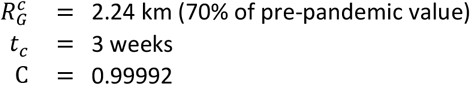

This coherent phase at optimal values becomes evident when we compare *R*_*G*_(*t*) with both *d*(*t*) and Δ*d*(*t*) as shown in Fig. 4, summarizing the main result of this work.

**Fig. 4.**
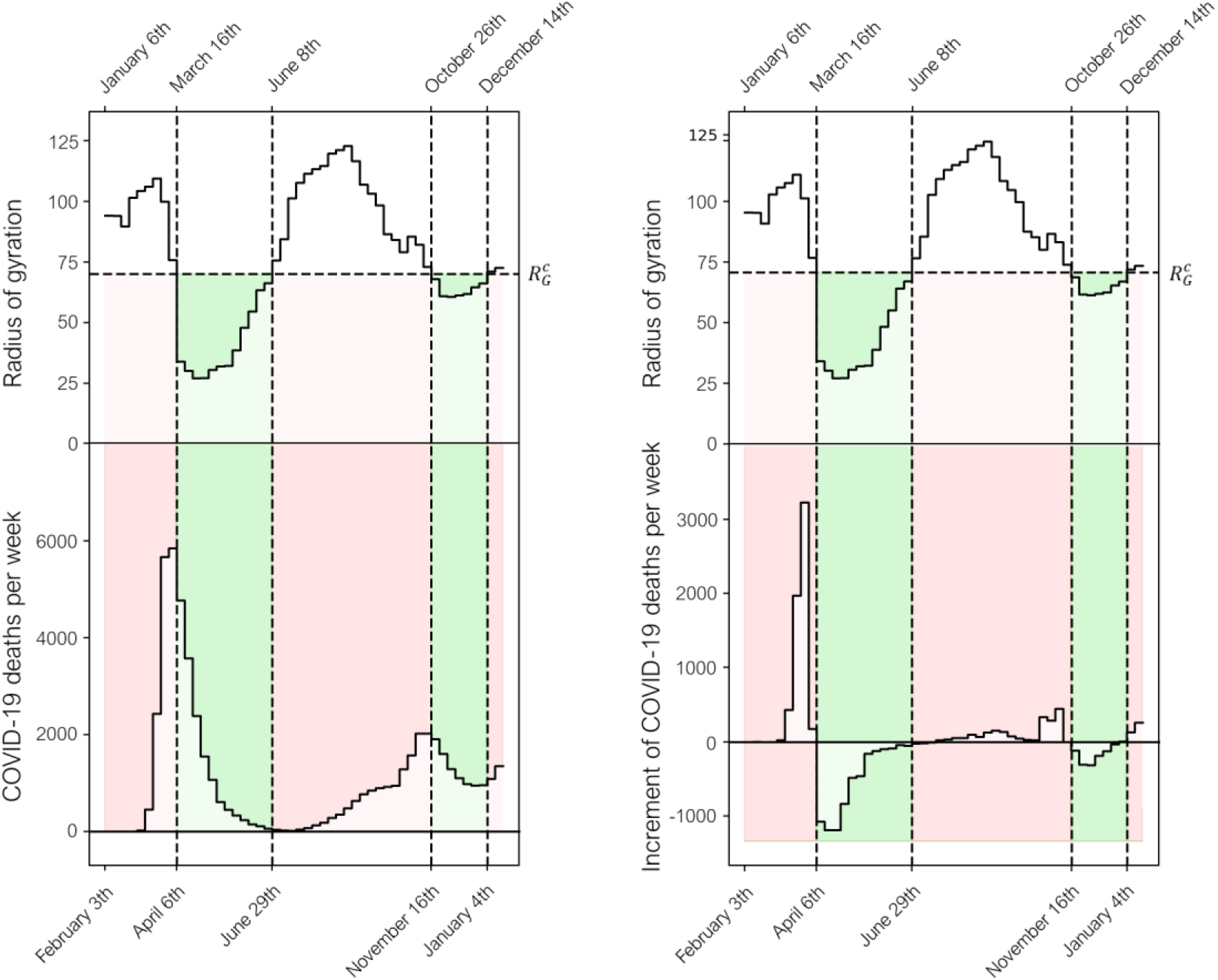
Left: Radius of gyration *R*_*G*_(*t*) compared with the number of COVID-19 deaths per week *d*(*t*) with an offset of 3 weeks; when *R*_*G*_ is above/below the critical value 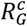, the number of deaths increases/decreases three weeks later (red/green areas). Right: Radius of gyration *R*_*G*_(*t*) compared with the weekly increment in the number of COVID-19 deaths per week Δ*d*(*t*) with an offset of 3 weeks; when *R*_*G*_ is above/below the critical value 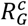, the increment in the number of deaths is positive/negative three weeks later (red/green areas).

This remarkable result shows that *R*_*G*_(*t*) has been an efficient predictor for the increment in the number of deaths per week for all 2020, with almost no exception (only two exceptions are found for values of Δ*d* very close to 0). Remarkably enough, the sign of Δ*d* has changed 4 times in 2020, fitting nicely with *R*_*G*_ crossing the critical value three weeks earlier. The offset of 3 weeks is compatible with empirical observations of periods between infection and death by COVID-19 [37].

The remarkable fit of the optimal values is also visible at the phase space diagrams of *R*_*G*_ − *d* and *R*_*G*_ − Δ*d*. Following both diagrams for all 2020 in Fig. 5, we find in the former case that the trajectory always moves up when is located at the right of the critical value, while it moves down when it crosses the critical value to its left. For the latter case we find that the trajectory always crosses quadrants passing by the (0,0) point: all points fall at the + + or the − − quadrant, and none at the + − or − + quadrants, as expected for two dynamical signals in phase.

**Fig. 5.**
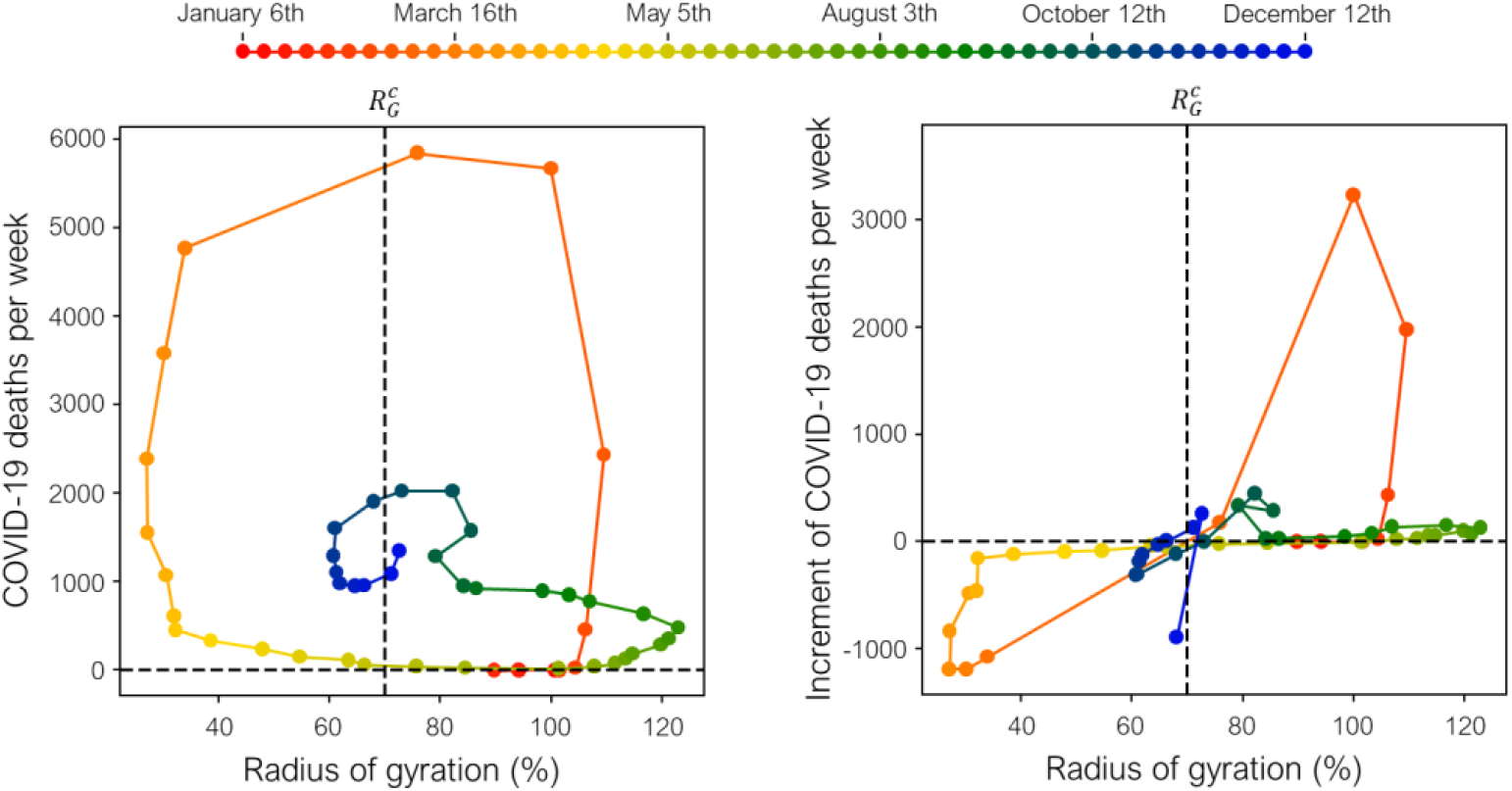
Left: Phase-space representation for *R*_*G*_ − *d* for optimal values. The trajectory always moves up al the right of the critical value, and always moves down at the left of the critical value. Right: Phase-space representation for *R*_*G*_ − Δ*d* for optimal values. The trajectory always cross the axes by(0,0), all points falling at + + or − − quadrants.

## 4. Discussion and conclusion

The radius of gyration is an efficient metric to describe the average mobility of the population in a particular region. When computed from mobile phone data as described in this work, it provides an accurate measurement of the effects of lockdowns and mobility restrictions, as well as changes in people’s behaviour. We show here that all relevant dates related with mobility measures or national holidays make an imprint on the weekly evolution of the median of the daily radius of gyration.

This remarkable accuracy helps to predict trends in the number of COVID-19 deaths, as soon as three weeks in advance. This offset of three weeks fits with the empirical periods between infection and death in the case of fatality by COVID-19, indicating that high values of radius of gyration are correlated with high infection rates, which ultimately leads to higher number of deaths weeks later. Thus, the radius of gyration is a valuable metric to monitor the effects of the mobility restrictions and to anticipate their impact.

An empirical critical threshold for the radius of gyration is observed in this work (70% of the pre-pandemic score) at which deaths increase after three weeks if the radius is above that value. The existence of that value indicates that partial lockdowns can be as efficient as total lockdowns, mitigating the collateral effects in the economy.

The mechanism underlying this critical value is unknown, and more research is needed. Massive mobile phone data contains rich information with high spatial and temporal resolution for a large number of users, allowing to aggregate and segment a representative sample of the whole population by a manyfold of parameters. We believe that a detailed segmentation of users by sociodemographic parameter as well as by region and mobility patterns will be instrumental in detecting and isolating the mechanism behind the critical value of the radius of gyration that correlates with the increment in the number of COVID-19 deaths.

## Data Availability

The information on mobility is available upon request with the permission of Kido Dynamics SA. The data on number of COVID-19 deaths can be downloaded from Ref. [32].

## Acknowledgements

The authors thank Jose Javier Ramasco from IFISC (UIB-CSIC) and Marta Gonzalez from University of Berkeley for useful discussions. This work received funding from the European Union’s Horizon 2020 research and innovation programme under grant agreement No 872249.

## Authors contribution

All authors verified data and results. All authors contributed equality to discussions and writing.

## Contact information

Dr. Alberto Hernando

